# Malaria Prevention Practices and Associated Factors among Households in Bube Town, Oromia Regional State, West Ethiopia

**DOI:** 10.64898/2026.01.15.26344173

**Authors:** Alemayehu Babusha Wega, Tefera Darge Beyene, Abebe Dukessa Dubiwak

## Abstract

**Background:** Malaria is a major health issue in Ethiopia, particularly in rural and semi-urban communities. The approaches used to combat malaria transmission in Ethiopia emphasize on vector control methods. However, the effectiveness of malaria prevention efforts may be influenced by difference in social and demographic factors, population’s knowledge and practices of malaria prevention. This study aimed to assess malaria prevention practices and associated factors among households in Bube Town, Oromia Regional State, Western Ethiopia.

**Objective:** To assess malaria prevention practices and associated factors among households in Bube Town, Oromia Regional State, West Ethiopia.

**Method:** A community-based cross-sectional study design was employed, with data collected via a questionnaire administered by interviewers. A systematic sampling technique was implemented during data collection, and the data were analyzed using SPSS version 25 after being entered into EpiData version 3.1 software. Binary logistic regression statistical analysis was done to determine association between dependent and outcome variables. Factors with P < 0.25 in the bivariate analysis were included in the multivariate regression analysis and eventually variables with a P-value < 0.05 at a 95% CI were considered to have a significant association with malaria prevention practices.

**Results:** The prevalence of good malaria prevention practices was 62.7% (95% CI: 58.0%–66.0%). Associated factors for good malaria prevention practices include higher age (above 25 years old), female sex (AOR=1.7, 95% CI: 1.035-2.295), knowledge of malaria breeding sites (AOR=3.7, 95% CI: 1.482-9.235), and knowing that malaria is preventable (AOR=4.014, 95% CI: 1.502-10.71).

**Conclusion:** More than half of Bube town households had good malaria prevention practices. Good practices were associated with being older, female, knowing where malaria breeds, and knowing that malaria is preventable. Raising community awareness and developing targeted interventions for males and young family members are crucial to advancing malaria prevention strategies.

## Background

Malaria is caused by species of the plasmodium parasite, namely P. falciparum, P. vivax, P. malariae, and P. ovale, and is transmitted by female anopheles mosquitoes (1). Although malaria can affect anyone, the groups most at risk of severe outcomes include pregnant women, infants, individuals with HIV/AIDS, and children under five years old (2, 3). If all preventative measures are implemented effectively, malaria can be both prevented and treated (4). The World Health Organization advocates for a set of prevention techniques aimed at decreasing the morbidity and mortality linked to the disease in regions where malaria is endemic (5). Strategies for controlling malaria have primarily focused on diminishing its burden through the use of insecticide-treated mosquito nets (ITNs), particularly long-lasting insecticidal nets (LLINs), and indoor residual spraying (IRS) (6). Educating communities about malaria prevention represents one of the strategies that can lower the risk of infection and related illnesses by promoting effective behavior change communication to alter attitudes towards prevention methods (7, 8).

The World Health Organization (WHO) estimated 263 million cases of malaria and 597,000 malaria-related deaths globally by 2023, with 94% of cases and 95% of deaths occurring in Africa (8, 9, 10). Malaria constitutes a major health challenge faced by African nations and significantly obstructs their socio-economic development, influencing both public health and national wealth; thus, it is regarded as both a disease and a source of poverty. The direct costs of malaria encompass both personal and governmental expenses related to its prevention and treatment, whereas indirect costs include lost productivity or income due to illness or mortality (11, 12). As malaria remains the most prevalent communicable disease in Ethiopian healthcare facilities, enhancing disease control efforts in malaria-prone areas of the country necessitates an integrated approach that incorporates community knowledge and practice. By 2018, the World Health Organization (WHO) suggested that approximately 72% of African households should have at least one insecticide-treated net (ITN), with 57% of the population being able to access one. Furthermore, it was recommended that 40% of the population live in households that have enough ITNs for all family members (3).

The study findings suggest that factors such as place of residence, education level, wealth, occupation, and cultural beliefs influence malaria prevention methods. These factors shape how communities adopt preventive and control strategies, and the effectiveness of these strategies may be hampered if community practices are not considered (13). Preferences and choices differ among communities (14, 15), and programs struggling to incorporate local malaria prevention practices have faced challenges in achieving long-term control (16). Despite the existence of effective preventive measures such as ITNs, IRS, and environmental management, their usage varies due to behavioral, socioeconomic, and logistical differences across regions. Bube Town in West Ethiopia, with both rural and urban characteristics, is a malaria-prone area with no existing studies evaluating malaria prevention practices and their related factors. This study aimed to examine malaria prevention practices and their determinants among households in Bube Town, Oromia Regional State, Western Ethiopia, in 2024.

## Methods and Materials

### Study Design

A community-based cross-sectional study design was used.

### Study Area

The study was conducted in Bube Town, situated in the Nole Kaba District, a part of the West Welega Zone, within the Oromia Region of Ethiopia. Nole Kaba shares borders with the Ilubabor Zone to the south, Kelem Welega Zone to the west, Yubdo to the northwest and Haru to the northeast. Bube town is approximately 411 km from Addis Ababa and 50 km from the zonal city. The town consists of two kebeles: Bube 01 and Bube 02, with a total population of 6,098 and 2,020 households, respectively.

### Study Period

The study was conducted from May 1 to June 30, 2024.

### Population

The source population comprised all households in Bube town, while the study population consisted of all selected households within the town.

### Inclusion and exclusion criteria

Individuals aged 18 years and above from the selected households who lived in the town for at least six months were included in the study. On the other hand, respondents from households who were unable to communicate due to impairment or serious illness were excluded from the study.

### Sample Size Determination and Sampling Technique

To calculate our sample size, we applied the single population proportion formula using n= (Z1-α/2)2 P (1-P)/d2, using P: 57.6% proportion of the population who drain stagnant water to prevent malaria, which gave the largest sample size among previous studies (13). We also applied the population correction formula nf = n / (1 + n / N) with a 10% non-response rate, resulting in nf = 343 participants. Considering a 10% non-response rate, our final sample size was approximately 377.

### Sampling Procedure

Households were selected using a systematic sampling approach. There were 2,020 households in the town of Bube. Thus, K = 2020 / 377 = 5.35, which was rounded to 5. The initial household was selected through a lottery method, followed by every 5th household thereafter.

### Data Collection Procedures

Interviewer-administered semi-structured questionnaires were used for data collection.

Five BSc nurses with prior experience in data collection and multilingual abilities were assigned to collect the data. Prior to the data collection period, the data collectors were trained to meet the study objectives. Throughout the data collection period, regular follow-ups and supervision were provided by the supervisor and principal investigator.

### Operational definitions

#### Attitude towards malaria

Attitudes were assessed using 13 positive and negative statements. The total score was calculated by summing the responses. A score of ≥23 indicated a positive attitude, whereas a score of ≤22 indicated a negative attitude.

#### Malaria prevention practices (Good or poor practice)

Individuals employ various methods to shield themselves from malaria. A practice is considered effective if the total score reaches (7.64) or higher after adding the responses to the seven related questions.

#### Knowledgeable (good knowledge)

The respondents’ comprehension of the fundamental details, symptoms, transmission, and prevention of malaria was assessed. A comprehensive knowledge score was determined by summing the scores for each respondent across all questions, assigning ’1’ for correct answers and ’0’ for incorrect answers, with a score exceeding 26 indicating a knowledgeable individual.

### Data quality management and statistical analysis

### Data quality management and statistical analysis

To assure validity, a pretest was conducted outside the study area prior to the real data collection period. Every day during and after data collection, the primary investigator and supervisors verified the gathered data for accuracy and consistency. During the data entry and analysis process, the data were edited, verified, coded, classified and tabulated.

### Informed consent

The Institutional Review Board of Mettu University reviewed and approved the study protocol (Ref. No: RPG/105/2024) and ethical clearance was obtained. Subsequently, an official letter was obtained from the Health Science Research and Postgraduate Coordinating Office and delivered to the Kebeles’ municipal authorities and Nole Kaba Health Office. All participants provided signed informed consent prior to data collection after being fully informed of the study’s objectives and purpose. Participants’ confidentiality was ensured during and after data collection.

### Ethics approval and consent to participate

This study was approved by the Mettu University Health Science College Research and Ethics Committee. All participants provided free and informed consent.

## Result

### Socio-demographic characteristics

Data were collected from 377 households, with a 100% response rate for each household. The average age of the participants was 31 (± 8.8), with males accounting for the majority (56.49%). Of the 377 participants, (43.50%) were between the ages of 25 and 34; (78.51%) were married, and 85 (15.91%) were single (Table 1).

**Table 1:** Socio-demographic characteristics of the study participants in Bube town, 2024

| Variable | Frequency | Percent (%) |
| --- | --- | --- |
| **Sex** |  |  |
| Male | 213 | 56.49 |
| Female | 164 | 43.50 |
| **Age in years** |  |  |
| 18-24 | 87 | 23.07 |
| 25-34 | 164 | 43.50 |
| 35-44 | 97 | 25.73 |
| >=45 | 29 | 7.69 |
| **Religion** |  |  |
| Orthodox | 108 | 28.64 |
| Protestant | 205 | 54.37 |
| Catholic | 30 | 7.96 |
| Muslim | 34 | 9.02 |
| **Marital status** |  |  |
| Married | 296 | 78.51 |
| Single | 60 | 15.91 |
| Divorced | 21 | 5.57 |

### Practice of malaria prevention methods

Of the 377 households that had heard about malaria, 184 (48.8%) used insecticide-treated mosquito nets. A total of 260(69%) participants cleaned stagnant water near their houses, but 117 (31.03 %) households did not participate in community malaria control campaigns. When sick, 236(62.7%) always attended health institutions, whereas 141(37.3%) never did (Table2).

**Table 2:** Practice of Malaria Prevention Measures in Bube Town, 2024

| Malaria prevention questions | Never(0) | Sometimes(1) | Always(2) |
| --- | --- | --- | --- |
| N (%) | N (%) | N (%) |  |
| How often do you sleep in insecticide treated mosquito net? | 43(11.4) | 150(39.8) | 184(48.8) |
| How often do you visit health facility when you or your families fall sick? | 44(11.7%) | 97(25.6) | 236(62.7) |
| How often do you use anti-mosquito spray (IRS) in Your house? | 121(32.2) | 217(57.6) | 39(10.2) |
| How often do you screen windows? | 82(21.7) | 177(47.1) | 118(31.2) |
| How often do you cut bushes around your house? | 22(5.9) | 247(65.4) | 108(28.8) |
| How often do you drain stagnant water around your home? | 28(7.3) | 261(69.3) | 88(23.4) |
| How often do you participate in malaria prevention campaign? | 211(55.9) | 153(40.7) | 13(3.4) |

The community’s overall practice score was 141 (37.3%) for poor practices and 236 (62.7%) for good ones. The figure below shows malaria prevention measures among families in Bube Town based on participants who learned about malaria within a year (Figure 1).

**Figure 1:**
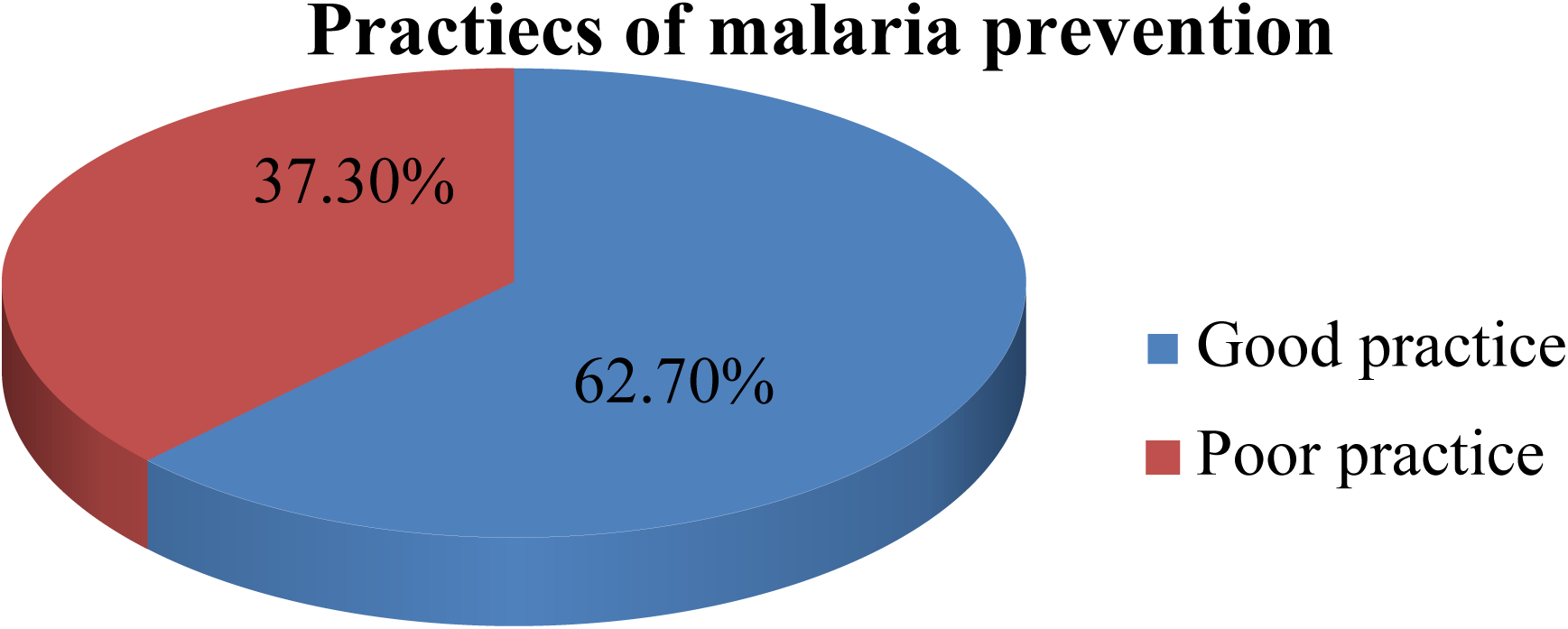
Malaria prevention practice level among households in Bube town, 2024.

### Knowledge about malaria prevention and control methods

Among the 377 individuals who had recently learned about malaria, nearly all (97.6%) recognized the primary symptoms, such as fever, chills, headache, loss of appetite, body aches, vomiting, and weakness. The majority (95.4%) agreed that mosquitoes are responsible for malaria, with most (93.7%) indicating that mosquito bites are the way malaria spreads. The majority of participants (90.7%) identified stagnant water, followed by swampy areas and waste, as potential mosquito breeding sites. Almost all (96.1%) of the participants were aware that mosquitoes bite, and over three-quarters (82.7%) mentioned that they bite at night.

Over nine out of ten (91.2%) respondents believed that malaria is preventable. ITN were identified as a malaria prevention strategy by 87.2% of respondents, followed by indoor residual house spraying (73.3%) and mosquito breeding source reduction (68.4%). Additionally, 65.1% of respondents understood that malaria primarily affects pregnant women and children (Table3).

**Table 3:** Knowledge on malaria prevention measures in Bube town, 2024

| Variable | Frequency | Percent % |
| --- | --- | --- |
| **Heard about malaria** |  |  |
| Yes | 374 | 99.3 |
| No | 3 |  |
| **Signs and symptoms of malaria** |  |  |
| Fever, chills, loss of appetite and joint pain | 370 | 98.3 |
| Vomiting and weakness | 229 | 60.7 |
| Urine color change | 151 | 40 |
| **Causes for malaria** |  |  |
| Mosquito | 362 | 96 |
| Other than mosquito | 217 | 57.6 |
| **Know breeding site** |  |  |
| Yes | 342 | 90.7 |
| No | 35 | 9.3 |
| **Breeding site** |  |  |
| Stagnant water and swampy areas | 310 | 82.3 |
| Waste materials | 213 | 56.5 |
| **Know biting time** |  |  |
| Yes | 362 | 96.1 |
| No | 15 | 3.9 |
| **Time of biting** |  |  |
| Day | 7 | 1.8 |
| Night | 318 | 82.7 |
| Any time | 58 | 15.5 |
| **Malaria preventable** |  |  |
| Yes | 344 | 91.2 |
| No | 33 | 8.8 |
| **Knowledge on preventive methods** |  |  |
| Use mosquito net | 328 | 87.2 |
| Indoor residual house spraying | 276 | 73.3 |
| Source reduction | 259 | 68.4 |
| Anti-malaria drugs | 232 | 61.5 |
| **Over all knowledge** |  |  |
| Good knowledge | 249 | 66 |
| Poor knowledge | 128 | 34 |

### Attitude of participants towards malaria

Participants’ attitudes towards malaria were categorized based on their attitude scores. A score of 23 or higher indicated a positive attitude, while a score lower than 22 signified a negative attitude. As a result, 62% of the study participants showed a positive attitude, while 38% demonstrated a negative attitude toward malaria, considering its susceptibility, severity, or threat level (Table4).

**Table 4:** Attitude towards malaria prevention measures in Bube town, 2024

| Attitude measuring questions | Agree<br>N (%) | Strongly<br>agree N (%) | Disagree<br>N (%) | Strongly<br>disagree<br>N (%) |
| --- | --- | --- | --- | --- |
| I think that Malaria is a life-threatening disease | 132(35.1) | 151(40) | 81(21.5) | 13(3.4) |
| Malaria is a communicable disease | 104(27.6) | 143(37.8) | 105(27.8) | 25(6.8) |
| I think the best way to prevent myself getting malaria is to avoid getting mosquito bites | 171(45.4) | 163(43.2) | 42(11.2) | 1 |
| I believe sleeping under a mosquito net during the night is one way to prevent myself getting Malaria | 146(38.8) | 211(56.1) | 17(4.4) | 3 |
| I am sure that self-treatment may dangers my health | 107(28.3) | 143(37.8) | 110(29.3) | 17(4.6) |
| In my opinion, children and pregnant women are at higher risk of Malaria | 143(38) | 198(52.7) | 26(6.8) | 10(2.4) |
| I think that one can't recover spontaneously from Malaria without any treatment | 80(21.2) | 171(45.4) | 100(26.6) | 26(6.8) |
| I think that malaria can't transmit through contact | 129(34.1) | 143(38) | 84(22.4) | 21(5.4) |
| I might be at a greater risk if not sleep inside ITN at night | 166(43.9) | 160(42.4) | 39(10.4) | 12(3.2) |
| I think that it is dangerous when malaria medicine is not taken completely | 130(34.4) | 176(46.8) | 60(15.9) | 10(2.9) |
| Seek for advice when get malaria | 209(55.6) | 141(37.3) | 23(6.1) | 4 |
| I seek treatment when get malaria | 195(51.7) | 158(42) | 18(4.9) | 6 |
| I think that I should have blood test if I have fever | 138(36.6) | 199(52.7) | 40(10.7) |  |
| Overall attitude |  |  |  |  |
| Positive attitude | 234(62) |  |  |  |
| Negative attitude | 143(38) |  |  |  |

### Factors Associated with malaria prevention practices among households

During the crude analysis, factors such as age, sex, occupation, marital status, religion, knowledge of mosquito breeding locations, knowledge of mosquito bite times, knowledge of how malaria spreads, and awareness of malaria prevention measures were considered for a multivariable logistic regression at p<0.25. Multivariable logistic regression revealed that age, sex, knowledge of mosquito breeding, and awareness of malaria prevention were significantly associated with good malaria prevention practices (p < 0.05). Compared to those between the ages of 18 and 24, those between the ages of 35 and 44 were 4.886 times more likely to practice good malaria prevention [AOR=4.886, 95% CI= (2.435, 9.805). Compared with males, females were more likely to take preventative action. Females were two times more likely to have good malaria prevention practice than males [AOR=1.74, 95%CI= (1.035, 2.295)].

Participants who knew where mosquitoes breed had 3.7 times higher chances of practicing good prevention measures than those who did not know [AOR=3.700, 95%CI= (1.482, 9.235)].The knowledge level of the study participants also influenced malaria prevention practices. Respondents who thought malaria was a preventable disease were 4.04 times more likely to use malaria prevention methods than those who thought it was not a preventable disease [AOR=4.04,95% CI= (1.502, 10.731) (Table 5).

**Table 5:** Bivariate and Multivariable Logistic Regression Analysis among households in Bube Town, Oromia Region, West Ethiopia, 2024

| Variable | Category | Practice |  | COR(95%CI) | AOR(95%CI) | P-value |
| --- | --- | --- | --- | --- | --- | --- |
|  |  | Good | Poor |  |  |  |
| Age | 18-24 | 33 | 54 |  | 1 |  |
|  | 25-34 | 130 | 48 | 4.452(2.646, 7.489) | 4.523(2.546, 8.032) | <0.08 |
|  | 35-44 | 56 | 30 | 2.948(1.646, 5.320) | 4.886(2.435, 9.805) | <0.001* |
|  | >=45 | 18 | 8 | 3.604(1.484, 8.749) | 5.812(2.029, 16.65) | 0.01* |
| Sex | Male | 120 | 96 | 106 | 1 |  |
|  | Female | 121 | 40 | 2.345(1.538, 3.575) | 1.74(1.035, 2.295) | P<0.001* |
| Marital status | Single | 50 | 68 | 75 | 1 |  |
|  | Married | 189 | 70 | 3.531(2.286, 5.457) | 6.93(3.615, 13.285) | 0.028 |
| occupation | Gov.t employee | 87 | 63 |  | 1 |  |
|  | Farmer | 28 | 11 | 1.619(0.180, 3.275) | 6.612(2.564, 17.05) | 0.06 |
|  | Private | 124 | 64 | 1.370(0.895, 2.095) | 3.484(1.838, 6.605) | 0.14 |
|  | employees |  |  |  |  |  |
| Know breeding site | No | 9 | 30 |  | 1 |  |
|  | yes | 231 | 107 | 6.721(3.094, 14.5) | 3.7(1.482, 9.235) | 0.005* |
| Is malaria preventable | No | 15 | 21 |  | 1 |  |
|  | Yes | 224 | 117 | 2.567(1.280, 5.146) | 4.014(1.502, 10.71) | 0.006* |

\*significant at P-Value <0.05, COR=crude odd ratio, AOR=adjusted odd ratio

## DISCUSSION

This study evaluated the factors influencing malaria prevention practices within the communities of Bube Town, located in the Oromia region of West Ethiopia. The findings revealed that 328 participants (80 %) were informed about malaria primarily through health facilities and health professionals rather than alternative sources. This figure exceeds the results of a study conducted in the Amhara region in Belesa (17). This difference may be due to the fact that the communities in our study area have limited access to other sources of information, such as mass media.

The majority, 379 (98.6%) of the study participants correctly identified fever, headache, chills, sweating, and malaise as the most common signs and symptoms of malaria. This is higher than the finding from Nigeria, where 65.2 percent of respondents indicated fever with shaking as a common malaria symptom (18). Botswana, where 88.7 percent of respondents had some basic understanding of malaria signs and symptoms(19), and another study conducted in Pawe District, North West Ethiopia, in which approximately 84.7% and 83% of the respondents stated fever and headache, respectively, as the most common primary symptoms associated with malaria(20). This study is consistent with a study conducted in Areka town, where fever, headache, chills, sweating, and malaise accounted for 94% of the most common signs and symptoms of malaria, and 94.4% in Shewa Robit town (21, 22).

This study shows that 91.2% of participant answered that malaria is preventable, and this This finding is consistent with a study conducted in Belesa and Woreta towns in Ethiopia(16,11).In this study the 62.7% of the study participants who always sought medical care when they or their family members were 29 sick. This is high when compared to two studies conducted in Ethiopian, 55.9% and 54.7 % (17, 23). Only 48.8 percent of participants who possessed ITNs always slept inside them; this finding is lower than studies conducted in Assosa town and outside, at Tumbi hospital in Tanzania, where In two studies, 58% and 76% of respondents used bed nets as the most common malaria protective measures, respectively(17,18). This is due to a lack of availability as well as a failure to employ available nets due to discomfort and high temperatures in the environment or it may be because of the lack of fear of mosquito bites or the belief that mosquitoes still bite while using ITNs. On the other hand only 57.2% households in this study sometimes sprayed by anti-mosquito spray, which is lower than other reports in north-eastern Ethiopia Shewa Robit town, which is 78.9%(21). This may be due to a program change made by the health care system from entire house-to-house spray to selective house spray. As a result, community healthcare providers might apply similar practices through health education initiatives. However, this is higher than two studies done in Gurage zone and Hawassa city administration whose 50% and 33.7% of households sometimes sprayed by the IRS (2, 24).

Approximately 70% of the study participants actively participated in the drainage of stagnant water and 65% cleaned bushes surrounding their houses. This is slightly lower than the study in southern region (24) but greater than the study conducted in Tanzania and Uganda (17, 20). This may be due to a lack of awareness of the negative consequences of the disease. Around 55.9% of participants never participated in malaria prevention campaigns, which is higher than the results of studies conducted in Ethiopia and Uganda (7, 16, 22). This might be the lack of strong leadership, participatory, and community-inclusive implementation programs. However, it is nearly similar to a study conducted in Hawassa city (2). The overall practice of malaria prevention through different strategies was much higher than in previous studies (22, 23). This can explained by intense preventive measures under taking by the government and non-governmental organizations through ITN distribution, availability of health Care facilities and the environment play major roles in malaria prevention programs.

In this study, the participants of ages above 24 are 4.45 times more likely to have good practice than age’s between18-24. This is in line with report from Thailand (25), but study in Uganda shows ages between 35-44 and 45+ were less likely to practice malaria prevention methods 30 compared to those ages between 18-24 (26). This might be because these age groups are more educated, have more access to health information than others, and are married; therefore, all household responsibilities belong to them, and they may receive bed nets for themselves and their children. On the other hand, participants who knew the breeding sites were 3.7 times more likely to practice prevention methods than those who did not. No similar study has been found, which may be because those who know the breeding sites of mosquitoes prevent mosquito breeding by reducing the source of breeding. Females were more likely to have good malaria prevention than males. This finding is supported by a study in West Belesa (27). This is because women are responsible for all home activities in this area; they take care for their children and take them to the hospital for treatment, they clean their homes as usual Ethiopian women do and clean the environment, and they have enough information on ITN use from health facilities during ANC follow-up, making them more responsible for malaria prevention.

Malaria is preventable, and those who know this have four times better malaria prevention practices than those who do not know this. Other studies in Ethiopia and other African countries have also shown that good knowledge is positively related to good malaria prevention practices (27, 28). This could be justified as participants who know that malaria is preventable also know the modes of transmission, risk factors, seriousness of the disease, and mechanisms of prevention, which may increase the probability of practicing preventive activities. Individuals who know that malaria is preventable have good motives for prevention.

### Limitations of the study

Our study has certain limitations; initially, we could not establish causality due to the cross sectional design of the study, and there may have been recall bias related to malaria prevention practices.

## Conclusion

Almost two-thirds of households in the study area follow recommended preventive measures, as evidenced by the prevalence of good malaria prevention practices, which was 62.7%. Being older than 25 years, being female, knowing where malaria breeds, and knowing that malaria is preventable were factors strongly associated with good malaria control practices. These findings suggest that health education and awareness campaigns targeting males and younger age groups could improve efforts to prevent malaria.

## Ethical consideration

The Institutional Review Board of Mettu University evaluated and approved the study protocol (Ref.No: RPG/206/2024), and ethical clearance was granted. Subsequently, a formal letter was requested from the Research and Postgraduate Coordinating Office of Health Science and submitted to the Nole Kaba Health office and local authorities. All procedures were executed in accordance with the applicable guidelines and regulations. Following a comprehensive explanation of the study’s aims and objectives, written informed consent was secured from all participants prior to the commencement of data collection. All data collected throughout the study was maintained in a confidential manner. The study was conducted in accordance with the Declaration of Helsinki.

## Ethics approval and consent to participate

This research got ethical approval by the Research and Ethics Committee of the Health Science College at Mettu University. All individuals involved in the study participated voluntarily and gave their informed consent.

## Consent for publication

Not applicable

## Availability of data and materials

The data sets employed and/or examined in this study can be obtained from the corresponding author upon a reasonable request.

## Competing interests

The authors declare that they have no competing interests

## Funding

The authors declare that no funds were received for this study.

## Authors’ Contributions

Mr.Alemayeu Babusha wrote the draft and performed an analysis

Mr. Tefera Darge performed wrote methodology

Mr. Abebe Dukessa performed data entry and edited the manuscript.

All authors read and approved the final manuscript

## Acknowledgments

We extend our sincere thanks to the Research Affairs Directorate of Mettu University and the Postgraduate Coordinating Office of the College of Health Sciences at Mettu University for their invaluable information and support. We also express our profound appreciation to the study participants and data collectors, as this research would not have been possible without their collaboration.

## List of abbreviations

ANC: Antenatal Care
AIDS: Acquired Immuno Deficiency Syndrome
HIV: Human Immuno Deficiency Virus
IRS: Indoor residual spray
ITN: Insecticide-treated nets
LLINs: Long-acting insecticide-treated nets
NGOs: Non-governmental organization
PMI: President’s malaria initiative
WHO: World Health Organization

## Notes

### Competing Interest Statement

The authors have declared no competing interest.

### Funding Statement

There is no funding received for this study

